# “Inhaled Nitric Oxide in Neonates with Pulmonary Hypertension in Amazonas, Brazil: Physiological Improvement Versus Impact on Relevant Clinical Outcomes”

**DOI:** 10.1101/2025.08.19.25333602

**Authors:** Rodrigo Duarte Ferreira, Cristina Helena Faleiros Ferreira, Walusa Assad Gonçalves-Ferri

## Abstract

**Background:** Inhaled nitric oxide (iNO) is a standard treatment for neonatal pulmonary hypertension in high-resource settings. Yet, its efficacy and cost-effectiveness in low- and middle-income countries (LMICs) remain underexplored. This study aimed to evaluate the impact of iNO on neonatal outcomes within a resource-limited Neonatal Intensive Care Unit (NICU) setting in Manaus, Brazil, to inform public health strategies.

**Methods:** We conducted a multicenter, quasi-experimental study employing a historical control design. We compared outcomes in 12 prospective neonates receiving iNO for persistent pulmonary hypertension of the newborn (PPHN) secondary to perinatal asphyxia (March-August 2018) with 12 historical controls (December 2015-December 2016). Participants were at a gestational age of more than 34 weeks with echocardiographic evidence of PPHN. Main outcomes included oxygenation parameters, mortality, and length of hospital stay.

**Results:** The prospective group demonstrated significant acute improvement in all key oxygenation parameters following initiation of iNO (p < 0.01 for PO2, O2 saturation, PO2/FiO2, and Oxygenation Index). However, iNO did not significantly reduce overall mortality (16.6% vs. 0%, p = 0.48) or NICU length of stay (21.3 vs. 13.2 days, p = 0.09). Notably, total hospital length of stay was significantly longer in the iNO group (37.1 vs. 23.08 days, p=0.03), with deaths primarily linked to systemic complications.

**Conclusion:** Although iNO acutely improves oxygenation in neonates with PPHN in this resource-limited setting, these physiological benefits did not result in reduced mortality or shorter NICU stays and were associated with increased overall hospitalization. The findings indicate that iNO sustains critically ill neonates who subsequently require extended care. Effective implementation of costly interventions in LMICs requires a comprehensive supportive infrastructure. Further context-specific research is crucial for informing resource allocation and enhancing neonatal care.v

## Introduction

Despite nitric oxide (NO) being a recommended treatment for neonatal pathologies involving pulmonary hypertension since the 1990s [1], which provides rapid and significant increases in preductal oxygen saturation and leads to the reversal of hypoxemia, some neonatal units worldwide still lack access to the gas, often due to economic reasons. [2]

Mortality has decreased in most units where NO has been introduced. However, the experience in developing countries has yielded different results. [2, 3] Implementing this therapy in healthcare units is expensive, and its use can lead to unjustified increases in healthcare costs. [4,5]

Limited comprehensive recent data on the long-term impact and cost-effectiveness of NO in resource-limited settings are available [3]. Further research is required to determine whether infants in developing countries experience clinical outcomes equivalent to those in developed countries when treated with inhaled nitric oxide. Such research is necessary to guide evidence-based resource allocation and the development of clinical guidelines in low- and middle-income countries (LMICs).

Infants in developing countries face a significant knowledge gap regarding optimal clinical interventions, as most research has been conducted in developed countries. Addressing this gap requires urgent promotion and support of clinical research in low- and middle-income countries (LMICs) to identify effective strategies for improving neonatal outcomes. Without such research, healthcare resources may be allocated to costly and ineffective treatments.

A multicenter study was designed to evaluate the implementation of nitric oxide therapy in Manaus City, Amazonas state, Brazil. The study aims to assess the impact of nitric oxide supply in neonatal units and to generate evidence regarding the feasibility and necessity of expanding this intervention to additional hospitals within the state of Amazonas.

## Methods

We conducted a multicenter, quasi-experimental study employing a historical control design on NICUs in Manaus, Amazonas State, Brazil, making NO available and promoting its use through guidelines. We evaluated the impact on neonatal outcomes. These ICUs are all tertiary complexity, public, and attend to outborn and inborn patients.

Six public tertiary maternity hospitals in Manaus were included, and data were collected from March to August 2018. We compared our results with historical controls of infants born between December 2015 and December 2016, using the same eligibility criteria. All hospitals involved in this study were tertiary-level NICUs.

Patients > 34 weeks of gestational age diagnosed with pulmonary hypertension (PH) based on echocardiographic evidence of increased pulmonary resistance and clinical signs [oxygenation index (OI) ≥ 25, severe hypoxemic respiratory failure, post-ductal PaO2 ≤ 60 mmHg with FiO2 at 100%, difference in pre- and post-ductal PaO2 or SatO2 > 20 mmHg or 5% (provided that SatO2 is between 70 and 95%), respectively, and/or two or more episodes of SatO2 falling below 85% in 12 hours] were included.

Patients who did not present with oxygenation disorders, complex congenital heart diseases, pulmonary malformations, pulmonary hypoplasia, congenital diaphragmatic hernia, or a lack of medical record data were excluded.

All staff members of the six neonatal units received training from the investigators and followed the guidelines of the Brazilian Health Ministry.[6]

Umbilical artery catheterization was performed to monitor blood gas levels through arterial blood gas analysis. Transfontanellar ultrasonography was performed during the first 72 h. NO (nitric oxide) and NO2 (nitrogen dioxide) levels, methemoglobinemia, and blood gas levels were continuously monitored. Blood clotting tests (PT and APTT) were performed every 24 h.

Data were expressed as means and standard deviations, medians and total variations, or numbers and proportions (percentages) according to the variable type. The two groups were compared using a non-parametric Wilcoxon test (for paired or unpaired data) or Fisher’s exact test, as appropriate. The adopted significance level was 5%.

Given the inherent rarity of persistent pulmonary hypertension of the newborn (PPHN) and the significant logistical and ethical constraints prevalent in this resource-limited setting, a formal sample size calculation was not performed. The ethical imperative primarily drove this decision to provide timely care, and the practical challenges of recruiting a large, prospectively defined cohort for a rare condition within a limited timeframe.

All stakeholders followed the gas usage guidelines when implementing the NO use. All patients in the NO group underwent echocardiography within the first 24 h. The NICU teams assembled and correctly installed devices and circuits for NO use without significant difficulties. No gas shortages are observed.

This project was approved by the Adriano Jorge Hospital Foundation Research Ethics Committee (11/21/2017, CAE 2·475·215). The parents’ consent was obtained.

## Results

During the prospective study period (March to August 2018), 12 patients were selected because of PPH (Persistent Pulmonary Hypertension) and neonatal asphyxia, with no patients excluded. In the retrospective group (December 2015 to December 2016), 23 were selected due to PPH and neonatal asphyxia, 11 were excluded due to insufficient medical record data to meet the inclusion criteria, and 12 were included. No significant differences were observed between groups (Figure 1).

**Figure 1.**
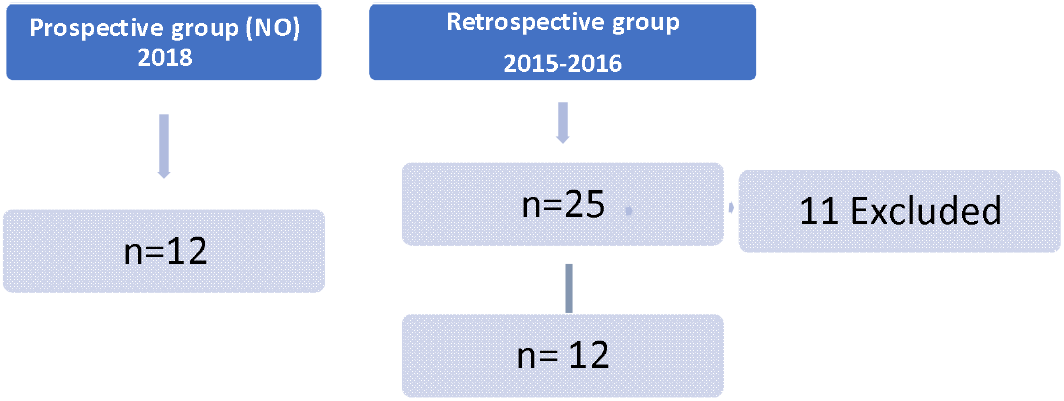
Study design.

The results regarding demographic and clinical variables between prospective and retrospective groups are presented in Table 1.

**Table 1:** Comparison of Demographic and Clinical Variables Between Prospective and Retrospective Groups.

| Variables |  | Prospective group (n=12) |  | Retrospective group (n=12) |  |
| --- | --- | --- | --- | --- | --- |
|  | N (%) | Mean ± SD | N (%) | Mean ± SD | p-value |
| Male | 6 (50) | - | 7 (58.3) | - | 1 |
| Weight (kg) | 12 (100) | 3.475 ± 1.20 | 12 (100) | 3.475 ± 1.04 | 0.89 |
| Vaginal | 6 (50) | - | 4 (33.3) | - | 0.68 |
| Gestational Age (weeks) | 12 (100) | 38.25 ± 2.86 | 12 (100) | 38.92 ± 1.68 | 1 |
| MAS* | 8 (66.7) | - | 9 (75) | - | 0.65 |
| Days in NICU |  | 21.3 (± 10.64) |  |  |  |
| Total Hospital Stay (days) |  | 37.1 (± 15.77) |  |  |  |
| Invasive Mechanical Ventilation (days) |  | 11.7 (± 6.53) |  |  |  |
| Death | 2 (16.6) | - | 0 | - | 0.48 |

\*MAS: Meconium Aspiration Syndrome

The mean number of hospitalization days in the prospective and retrospective groups was 37,1 and 23,08 days, respectively. Regarding NO introduction, the patients used gas for an average of 6 days (SD ± 3,24) and 25 parts per million doses (SD± 14,6).

The data about gasometric values are presented in Table 2.

**Table 2:** Gas blood values characteristics before and after the introduction of NO in the prospective group.

| Variables | Before NO |  |  | After NO |  |  | p* |
| --- | --- | --- | --- | --- | --- | --- | --- |
|  | Mean (SD) | Minimum | Maximum | Mean (SD) | Minimum | Maximum |  |
| pH | 7,11 (±0,17) | 6,82 | 7,29 | 7,22 (±0,16) | 6,88 | 7,42 | 0,01 |
| PO2 | 42,3 (±9,7) | 27,4 | 60 | 95,1(±46,62) | 19,4 | 167 | <0.01 |
| PaCO2 | 76,7 (±34,7) | 32,7 | 172,2 | 63,49 (±35,9) | 24,5 | 138,8 | 0.04 |
| Bicarbonate | 18,4 (±6,32) | 6,2 | 28,2 | 20,18 (±4,9) | 12,5 | 31,1 | 0.72 |
| Base Excess(mmMol/L) | -4,81(±7,48) | -23 | 7,9 | -3,28 (±5,28) | -14 | 8,5 | 0.75 |
| O2 Saturation | 68,74(±20,1) | 33,6 | 88,6 | 87,5 (±23,24) | 23 | 98,9 | <0.01 |
| PO2/FiO2 | 42,28 (±9,7) | 27,4 | 60 | 95,03 (±46,6) | 19,4 | 167 | <0.01 |
| OI* | 34,5 (±12,4) | 25 | 59 | 21,01(±14,83) | 7 | 56 | <0.01 |

\* OI Oxygenation index
\* \* Wilcoxon test non-parametric

We found a significant improvement in blood gas parameters (PaO2 and PO2/FiO2) after introducing NO (Table 1). However, two patients died in the prospective group: one due to septic shock at 48 h of life, who did not respond to NO, and the second at six days of life due to late neonatal sepsis. The average length of stay in the NICU in the prospective and retrospective groups was not significantly different (21,3 days vs 13,2 days, respectively). Regarding mortality, no statistically significant differences were observed between groups.

## Discussion

This study evaluated the effects of inhaled nitric oxide (iNO) therapy on physiological and clinical outcomes in neonates with persistent pulmonary hypertension of the newborn (PPHN) due to perinatal asphyxia in Manus NICUs, a resource-limited setting. We observed a significant acute improvement in oxygenation parameters, including the Oxygenation Index, in the prospective group after initiation of iNO (all p < 0.01). This aligns with iNO’s known role as a selective pulmonary vasodilator that improves ventilation-perfusion matching. The average iNO duration (6 days) and dose (25 ppm) were consistent with international guidelines, supporting the immediate effectiveness of iNO in reversing pulmonary vasoconstriction and stabilizing critically ill neonates [3-6].

Despite these clear acute physiological benefits, our study did not demonstrate a statistically significant reduction in overall mortality. This finding aligns with large randomized controlled trials (RCTs) suggesting that iNO, in term or near-term neonates with PPHN, primarily reduces the need for extracorporeal membrane oxygenation (ECMO) rather than directly impacting mortality. In a meta-analysis, Barrington et al. (2017) reported that NO treatment reduced the incidence of death and the need for ECMO [7]. However, only one study in that review was conducted in a developing country (Chile), and its focus was on oxygenation rather than mortality or other morbidities [8].

Furthermore, deaths in our prospective cohort were linked to severe systemic complications, such as septic shock and late-onset neonatal sepsis, underscoring that iNO, while effective for pulmonary issues, does not mitigate risks stemming from underlying systemic conditions or nosocomial infections. This reinforces the necessity of a holistic therapeutic approach that extends beyond isolated pulmonary intervention. Articles on the use of iNO in developing countries have frequently evaluated its immediate effect on blood gas values (oxygenation), showing positive results. Nevertheless, impacts on mortality and morbidity were often unrelated, leaving the overall impact of iNO in resource-limited settings unclear [9-14].

Regarding length of stay and ventilation, no statistically significant differences were observed in NICU length of stay (21.3 days vs. 13.2 days, p = 0.09) or invasive mechanical ventilation days (11.7 days vs. 8.67 days, p = 0.33). However, the total hospital length of stay was significantly longer in the prospective group (37.1 days vs. 23.08 days, p = 0.03). This outcome may suggest that iNO enabled the survival of more critically ill neonates who, without this intervention, might have succumbed earlier.

The absence of reduced ventilation duration suggests that although iNO improves oxygenation and may aid extubation, it does not significantly decrease the need for mechanical ventilatory support in complex cases where pulmonary recovery or resolution of organ dysfunctions are limiting factors. Other treatments effective in high-resource settings may not yield similar results in developing countries due to overcrowding, staff shortages, and supply limitations.

Similarly, more data are needed on the effectiveness of other neonatal treatments in LMICs; for instance, a recent review identified only three out of 27 studies on infection control measures conducted in resource-limited settings, underscoring the evidence gap for these interventions [16-17]. Additionally, a critical shortage of healthcare workers with adequate competencies, as well as systemic challenges such as poor education, limited access to evidence-based guidelines, and inadequate work environments, contribute to suboptimal care in LMICs [21, 28, 29].

The interpretation of these results must consider the methodological limitations inherent to our study design. As a quasi-experimental study utilizing a historical control group, a pragmatic choice given the rarity of PPHN and the ethical/logistical challenges of conducting an RCT in a resource-limited environment, it is susceptible to selection biases and confounding variables. While efforts were undertaken to ensure the comparability of measurable baseline characteristics between the groups (e.g., weight, gestational age, sex, type of delivery, MAS -Meconium Aspiration Syndrome), these inherent temporal variations and the absence of concurrent control fundamentally limit our ability to establish definitive cause-and-effect conclusions regarding the isolated impact of iNO. Moreover, the small sample size (12 patients per group) is a significant limitation, particularly in detecting differences in low-incidence outcomes, such as mortality, which increases the risk of a Type II error. Consequently, our findings are best interpreted as suggestive and exploratory, with their generalizability primarily limited to contexts sharing similar characteristics and resource availability [15-29].

This study offers valuable insights into the clinical application of inhaled nitric oxide (iNO) in resource-limited settings, which are often underrepresented in the global medical literature. Data from high-resource environments may not be directly applicable, emphasizing the need for context-specific evidence to inform local clinical practice and policy. By addressing the evidence gap in low- and middle-income countries (LMICs), this research highlights the challenges and outcomes associated with implementing advanced therapies without a comprehensive support infrastructure. Publishing these findings supports further research and promotes evidence-based interventions tailored to LMICs.

## Conclusion

This study demonstrates that inhaled nitric oxide (iNO) improves oxygenation in neonates with persistent pulmonary hypertension of the newborn (PPHN), including in resource-limited settings. However, this improvement did not result in a significant reduction in mortality or a shorter duration of admission to the neonatal intensive care unit (NICU). To optimize outcomes associated with high-cost interventions such as iNO, neonatal units should implement standardized clinical management protocols and strengthen overall quality of care.

## Data Availability

All data produced in the present work are contained in the manuscript.

## List of abbreviations

NO: Nitric oxide
NICUs: Neonatal intensive care units
LMICs: Low-Middle Income Countries
NO2: nitrogen dioxide
PPH: Persistent Pulmonary Hypertension

## REFERÊNCIAS

1. Roberts, J. D. et al. Inhaled nitric oxide in persistent pulmonary hypertension of the newborn. Lancet, London, v. 340, n. 8823, p. 818–819, 3 out. 1992.

2. Nakwan, N. The practical challenges of diagnosis and treatment options in persistent pulmonary hypertension of the newborn: a developing country’s perspective. American Journal of Perinatology, [S. l.], 2018.

3. Bandiya, P.; Madappa, R.; Joshi, A.R. Etiology, diagnosis, and management of persistent pulmonary hypertension of the newborn in resource-limited settings. Clinics in Perinatology, Philadelphia, v. 51, n. 1, p. 237–252, Mar. 2024.

4. Ho, T. et al. Choosing wisely in newborn medicine: five opportunities to increase value. Pediatrics, Itasca, v. 136, n. 2, p. e482–e489, 2015.

5. Fuentes, C. K.; Vallejos, C.V.; Navarro, A.R. Costo efectividad y análisis de impacto presupuestario del óxido nítrico inhalatorio neonatal en un hospital, desde la perspectiva del sistema público de salud. Revista Chilena de Pediatría, Santiago, 2016.

6. BRASIL. Ministério da Saúde. Atenção à saúde do recém-nascido. In Brazil. Ministério da Saúde. Secretaria de Atenção à Saúde. Departamento de Ações Programáticas Estratégicas. Atenção à saúde do recém-nascido: guia para os profissionais de saúde. Brasília, DF: Ministério da Saúde, 2012. p. 70–81.

7. Barrington, K. J. et al. Nitric oxide for respiratory failure in infants born at or near term. Cochrane Database of Systematic Reviews, London, n. 1, CD000399, 2017.

8. GonzÁLez, A. et al. Randomized controlled trial of early compared with delayed use of inhaled nitric oxide in newborns with moderate respiratory failure and pulmonary hypertension. Journal of Perinatology, London, v. 30, n. 6, p. 420–424, Jun. 2010.

9. Abdallah, V. O. S. et al. Óxido nítrico inalatório no tratamento da hipertensão pulmonar persistente do recém-nascido. Revista Médica de Minas Gerais, Belo Horizonte, v. 22, n. 4, p. 374–379, 2012.

10. Cabral, J. E. B.; Belik, J. Persistent pulmonary hypertension of the newborn: recent advances in pathophysiology and treatment. Jornal de Pediatria, Rio de Janeiro, v. 89, n. 3, p. 226–242, maio/jun. 2013.

11. Lopes, J. M. A. et al. Óxido nítrico no tratamento da hipertensão pulmonar persistente do recém-nascido. Jornal de Pediatria, Rio de Janeiro, v. 72, n. 3, p. 133–138, maio/jun. 1996.

12. Coletti, K. et al. Randomized controlled trials of pulmonary vasodilator therapy adjunctive to inhaled nitric oxide for persistent pulmonary hypertension of the newborn: a systematic review. Clinics in Perinatology, Philadelphia, v. 51, n. 1, p. 253–269, Mar. 2024. DOI: 10.1016/j.clp.2023.11.009.

13. Kazi, S. et al. The utility of chest x-ray and lung ultrasound in the management of infants and children presenting with severe pneumonia in low- and middle-income countries: a pragmatic scoping review. Journal of Global Health, Edinburgh, v. 12, ID 10013, 23 dez. 2022. DOI: 10.7189/jogh.12.10013.

14. Muriuki, A. et al. On the road to universal coverage of postnatal care: considerations for a targeted postnatal care approach for at-risk mother-baby dyads in low-income and middle-income countries informed by consultation with global experts. BMJ Open, London, v. 12, n. 6, e058408, 14 jun. 2022. DOI: 10.1136/bmjopen-2021-058408.

15. Jury, I.; Thompson, K.; Hirst, J.E. A scoping review of maternal antibiotic prophylaxis in low- and middle-income countries: comparison to WHO recommendations for prevention and treatment of maternal peripartum infection. International Journal of Gynaecology and Obstetrics, Chichester, v. 155, n. 3, p. 319–330, dez. 2021. DOI: 10.1002/ijgo.13648.

16. Graham, W. J. et al. Cleaning neonatal units in low-resource settings: a hot topic in waiting? Review. Pediatric Infectious Disease Journal, Philadelphia, v. 40, n. 5S, p. S1–S4, 1 maio 2021.

17. Fitzgerald, F. C. et al. The impact of interventions to prevent neonatal healthcare-associated infections in low- and middle-income countries: a systematic review. Pediatric Infectious Disease Journal, Philadelphia, v. 41, n. 3S, p. S26–S35, 1 mar. 2022.

18. Cross, S. et al. An invisible workforce: the neglected role of cleaners in patient safety on maternity units. Global Health Action, Stockholm, v. 12, n. 1, 1480085, 2019.

19. Bolan, N. et al. Human resources for health-related challenges to ensuring quality newborn care in low- and middle-income countries: a scoping review. Global Health: Science and Practice, Baltimore, v. 9, n. 1, p. 160–176, 31 mar. 2021. DOI: 10.9745/GHSP-D-20-00362.

20. Javaid, A.; Syed, S. Infant nutrition in low- and middle-income countries. Clinics in Perinatology, Philadelphia, v. 49, n. 2, p. 475–484, Jun. 2022.

21. Debbag, R. et al. Are the first 1,000 days of life a neglected vital period to prevent the impact on maternal and infant morbimortality of infectious diseases in Latin America? Proceedings of a workshop of experts from the Latin American Pediatric Infectious Diseases Society, SLIPE. Frontiers in Pediatrics, Lausanne, v. 11, ID 1297177, 30 nov. 2023.

22. Barron-Garza, F. et al. Incidence of cerebral palsy, risk factors, and neuroimaging in northeast Mexico. Pediatric Neurology, New York, v. 143, p. 50–58, Jun. 2023.

23. Blanco, E. et al. Adverse pregnancy and perinatal outcomes in Latin America and the Caribbean: systematic review and meta-analysis. Revista Panamericana de Salud Pública, Washington, DC, v. 46, e21, 2 maio 2022.

24. Silva, J. C.; Zin, A.; Gilbert, C. Retinopathy of prematurity prevention, screening and treatment programs: progress in South America. Seminars in Perinatology, Philadelphia, v. 43, n. 6, p. 348–351, out. 2019.

25. Hazel, E. A. et al. Neonatal mortality risk of vulnerable newborns by fine stratum of gestational age and birthweight for 230,679 live births in nine low- and middle-income countries, 2000-2017. BJOG: An International Journal of Obstetrics & Gynaecology, Chichester, 16 Jan. 2024.

26. Garg, P. Do we need to use nitric oxide in preterm babies in developing countries? Ceylon Medical Journal, Colombo, v. 51, n. 1, p. 22–26, Mar. 2006.

